# Assessment of Geriatric Depression among Elderly Peoples Residing at Rural Area of Kheda District, Gujarat

**DOI:** 10.1101/2023.04.10.23288346

**Authors:** Vaibhavi Parmar, Kailash Nagar, Virendra Jain

**Affiliations:** Dinsha Patel College of Nursing, Nadiad

**Author notes:** **Corresponding Author:** - Mr. Kailash Nagar (Email Id). **Researcher:** Ms. Vaibhavi parmar (Email Id).

**Keywords:** (GDS) Geriatric Depression scale, Elderly, Rural area, world health organization

## Abstract

**Background:** Depression is a major mental health problem, which is yet to be recognized as an important public health challenge. About 322 million people affected with depression worldwide.

Depression among the elderly population in India has been recognized as one of the major public health problems with a prevalence of 8 to 22%. It causes significant suffering and accounts for 5.7% of the years lived with disability, to make matters worse, it has been reported that in many cultures and societies, deteriorating mental status, say it dementia or depression, has been perceived as a part of normal ageing.

**Aims and Objectives:** The aim of this study is to explore the magnitude and risk factors of depression in elderly people residing in rural area of Gujarat.

*Objective:* 1. To assess the level of depression among elderly people residing at rural area. 2. To find out Significant Association between the level of depression selected demographic variables among elderly people at rural area.

**Methodology:** *Design and Setting:* A cross sectional descriptive study design was used. Non probability convenient sampling technique was used to drawn samples. 100 elderly persons were screened. Geriatric depression scale (GDS) was used to assess depressive level. A self-rating questionnaire tool was used to collect data and some socio-demographics and clinical variables. Prior to data collection written setting permission obtain from authority of the village and written consent from was obtained from the participants. Data for study was collected by door to door visit in selected community. Chi square was employed to determine association of collected data. **Tool consists of following <u>Section A:</u>** Socio-demographic variables. **<u>Section B:</u>** Geriatric Depression Scale (GDS). The GDS questions are answered ‘yes’ or ‘no’, instead of a five-category response set. This simplicity enables the scale to be used with ill or moderately cognitively impaired individuals. The scale is commonly used as a routine part of a comprehensive geriatric assessment. One point is assigned to each answer and the cumulative score is rated on a scoring grid. The grid sets a range of 0–9 as ‘normal’, 10–19 as ‘mildly depressed’, and 20–30 as ‘severely depressed.

**Results:** Results showed that 61% was Male and 39% was Female. The majority of had 71% primary education. Regarding work majority 60% are labor, regarding financial dependency most 62% was independent, 35(35%) dependent. This study reveals that level of depression among geriatric peoples, 51 (51%) peoples had no depression, 33 (33%) having mild depression and 16 (16%) having severe depression level.

**Conclusion:** It is concluded that nearly 50% of geriatric peoples having no depression and mild depression is more prevalent among elderly in selected rural area of Gujarat. So majority of the elderly peoples were found normal.

## INTRODUCTION

Geriatric depression is a mental and emotional disorder affecting older adults. Geriatric depression scale is used to quantify the severity of depression. Empty nest syndrome (loneliness in elderly is common causative factor for depression) is a factor should be rectified by social skills training.^1^ Late onset bipolar disorder is rare. Depression is common among elderly.^2^ It is a mental health condition that can affect people of all ages. While it is normal to feel down sometimes, if you feel this way for 2 weeks or more, or your mood is affecting your ability to cope with everyday life, you may be experiencing depression.^**3**^ In elderly people, depression mainly affects those with chronic medical illnesses and cognitive impairment, causes suffering, family disruption, and disability, worsens the outcomes of many medical illnesses, and increases mortality.^4^

According to WHO, factors increasing depression risk in older adults include genetic, chronic disease and disability, pain, frustration with limitations in activities of daily living, personality traits (dependent, anxious or avoidant), adverse life events (separation, divorce, bereavement, poverty, social isolation) and lack of adequate social support. Many studies have demonstrated a relationship between depression and various socioeconomic variables such as advanced age, low education, poverty and manual occupation. Thus, an older adult patient suffering from depression often has a combination of psychological, physical and social needs.^5^

More than 1 in 10 older people, and more than 3 in 10 people living in residential aged-care, experience depression. It’s important to remember that not all older people become depressed, and just because you are older, you don’t need to accept that you will become depressed, or that your depression can’t be treated. Technical advances have facilitated the exploration of factors related to geriatric depression and have helped generate novel biological and psychosocial treatment approaches.^6^

Based on UNDESA’s 2019 report, the population aged over 65 years old would reach 703million and was predicted to double by 2050, that is, 1.5 billion in which the proportion of older adult is 16%. It means that 1 of 6 people in the world’s population is elderly.^7^ Depression is one of the most common illnesses worldwide, with more than 264 million people affected. Various studies in the elderly population have estimated the prevalence of depression across India, with results ranging from 6% to 62%. The objectives of this study were to estimate the prevalence of depression among the elderly population using a Geriatric Depression Scale (GDS) and to find out the association between various sociodemographic parameters and depression among elderly people.^8^ Depression or the occurrence of depressive symptomatology is a prominent condition amongst older people, with a significant impact on the well-being and quality of life. Many studies have demonstrated that the prevalence of depressive symptoms increases with age (Kennedy, 1996). Depressive symptoms not only have an important place as indicators of psychological well-being but are also recognized as significant predictors of functional health and longevity. Longitudinal studies demonstrate that increased depressive symptoms are significantly associated with increased difficulties with activities of daily living (Penninx *et al*., 1998). Community-based data indicate that older persons with major depressive disorders are at increased risk of mortality (Bruce, 1994).^9^

Attention guides thought and behaviour. Information that is attended becomes available to higher-order cognitive processes such as working memory and decision-making. In order to promote wellbeing, it is important that attention selects stimuli associated with rewarding outcomes (Anderson, 2013).^10^ Because of its devastating consequences, late life depression is an important public health problem. It is associated with increased risk of morbidity, increased risk of suicide, decreased physical, cognitive and social functioning, and greater self-neglect, all of which are in turn associated with increased mortality.^10^

## OBJECTIVE

1. To assess the level of depression among elderly people residing at rural area of Kheda district
2. To find out Significant Association between the level of depression selected demographic variables among elderly people at rural area of Kheda, district.

## MATERIALS AND METHODS

### Research Approach

Quantitative Research approach was used for the current study.

### Research Design

The design of the study will be used Non-Experimental Cross-Sectional Survey Research Design.

### Research Variables: There are two types of variables considered under the study as follows

#### 1. Research variables

In present study research variable is the level of depression among old age people.

#### 2. Demographic variables

Age, gender, religion, education, marital status, type of family, locality of house, no. of children, frequency of visit of children or relative, hobbies, financial support, Previous occupation, old age pension

### Sampling method

The sample of the study will be selected by using **<u>Non probability convenient sampling technique</u>**. According to inclusive criteria as well as availability of samples.

### Study population

In the present study, the target populations are old age people residing at old age residing at rural area of selected areas of Kheda district”.

### Study Sample

Geriatric peoples residing at rural area of selected areas of Kheda district”.

### Study Setting

PHC Pethai has been selected for the main study setting and data collect from the elderly people.

### Sample Size

100 geriatric people

## SAMPLE CRITERIA

### Inclusion criteria

1. Old age people who are aged 60year and above.
2. Old age people who are willing to participate in the study.
3. Old age people who are available at the time of study.

### Exclusion criteria

1. Those who are present at data collection.
2. Those who are seriously ill
3. Those who are not able to interact (hearing loss and language problems)

### Tool for Data Collection

<u>Section A:</u> Sociodemographic variables.

<u>Section B:</u> Geriatric Depression Scale.

## RESULTS

The above table 2.1 shows level of depression among geriatric peoples, 51 (51%) peoples had no depression, 33 (33%) having mild depression and 16 (16%) having severe depression level.

**TABLE–1:**
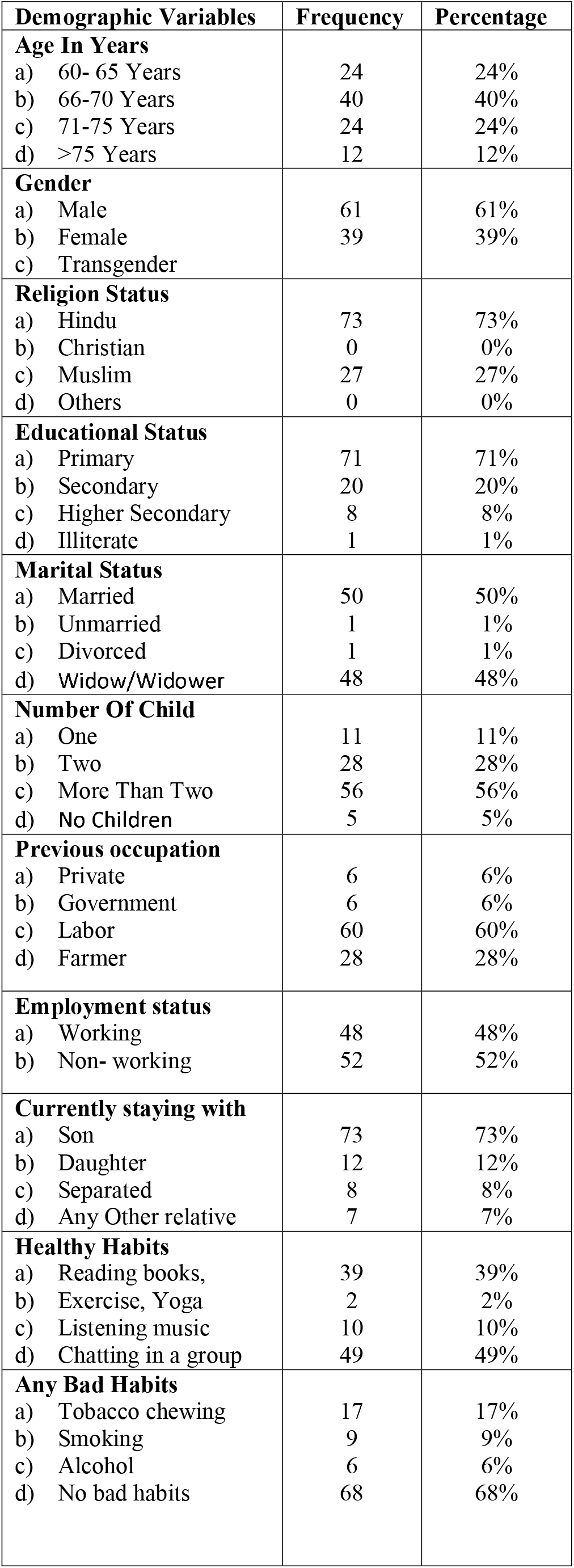
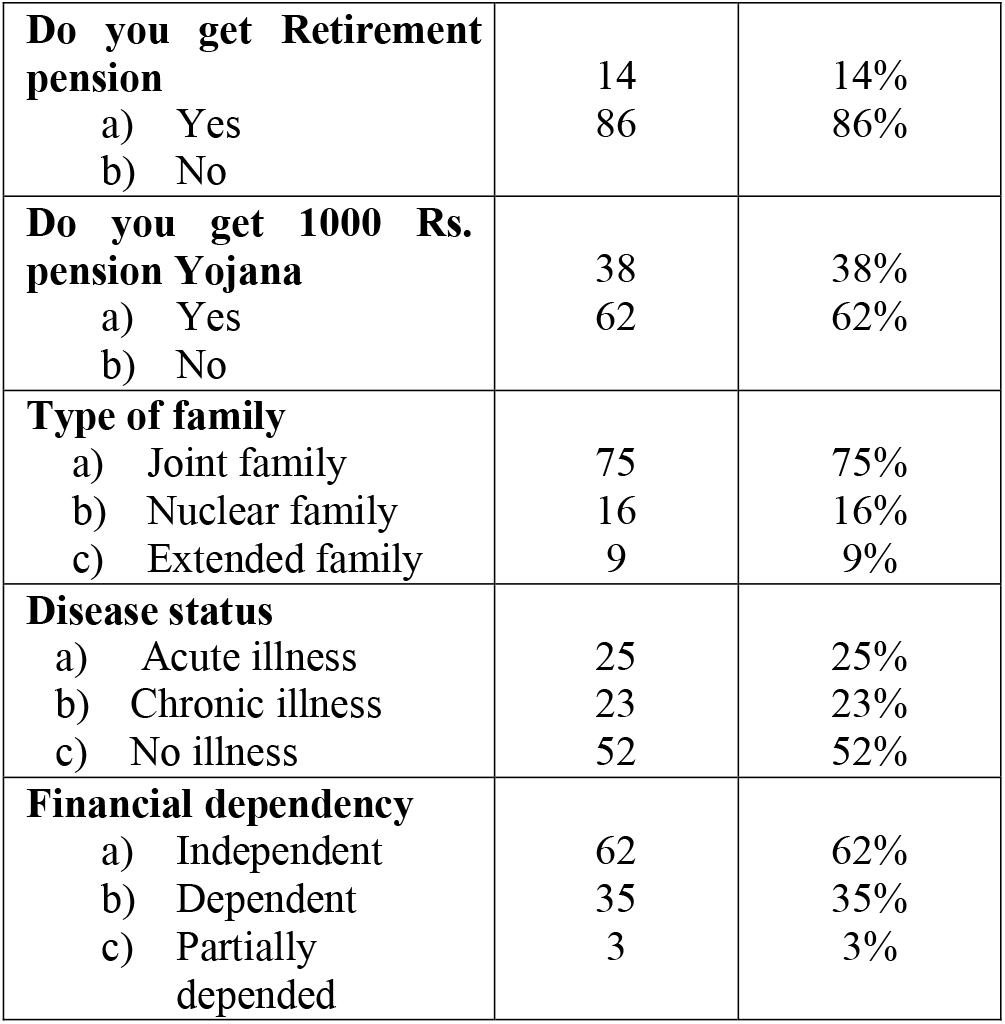
Socio-demographic variables of geriatric peoples residing at rural area of Kheda district N=100.

**TABLE–2.1:** Distribution of the geriatric Peoples according to Level the Depression.

| Level of Depression | Frequency (%) |
| --- | --- |
| No Depression | 51 (51%) |
| Mild Depression | 33 (33%) |
| Severe Depression | 16 (16%) |
| Total | 100 (100%) |

**Graph no.1.**
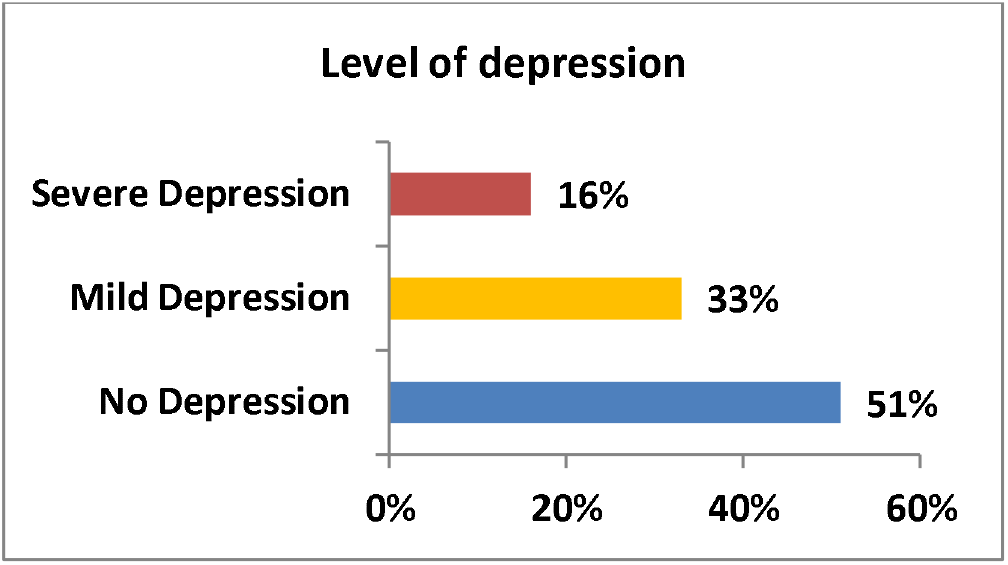
showing distribution according to level of depression among geriatric peoples.

The above table 2.2 shows depression score range was 1-24, mean score was 10.97, and standard deviation was 6.014

**TABLE–2.2:** Range, Mean, and Standard Deviation of depression among the geriatric peoples residing at rural area of Kheda district.

| Level of knowledge | Max. score | Range | Mean | SD |
| --- | --- | --- | --- | --- |
| Overall | 24 | 23 | 10.97 | 6.014 |

The table no.3.1 envisages the outcome of chi-square analysis being carried out to bring out the association between the mean difference level of depression among geriatric peoples living at rural area of Kheda district with their selected demographic variables, Age, Religion Status, Educational Status, Marital status, Number of Child, Occupation, habits, Pension etc. were accounted for determining the association with level of depression.

**TABLE–3.1:**
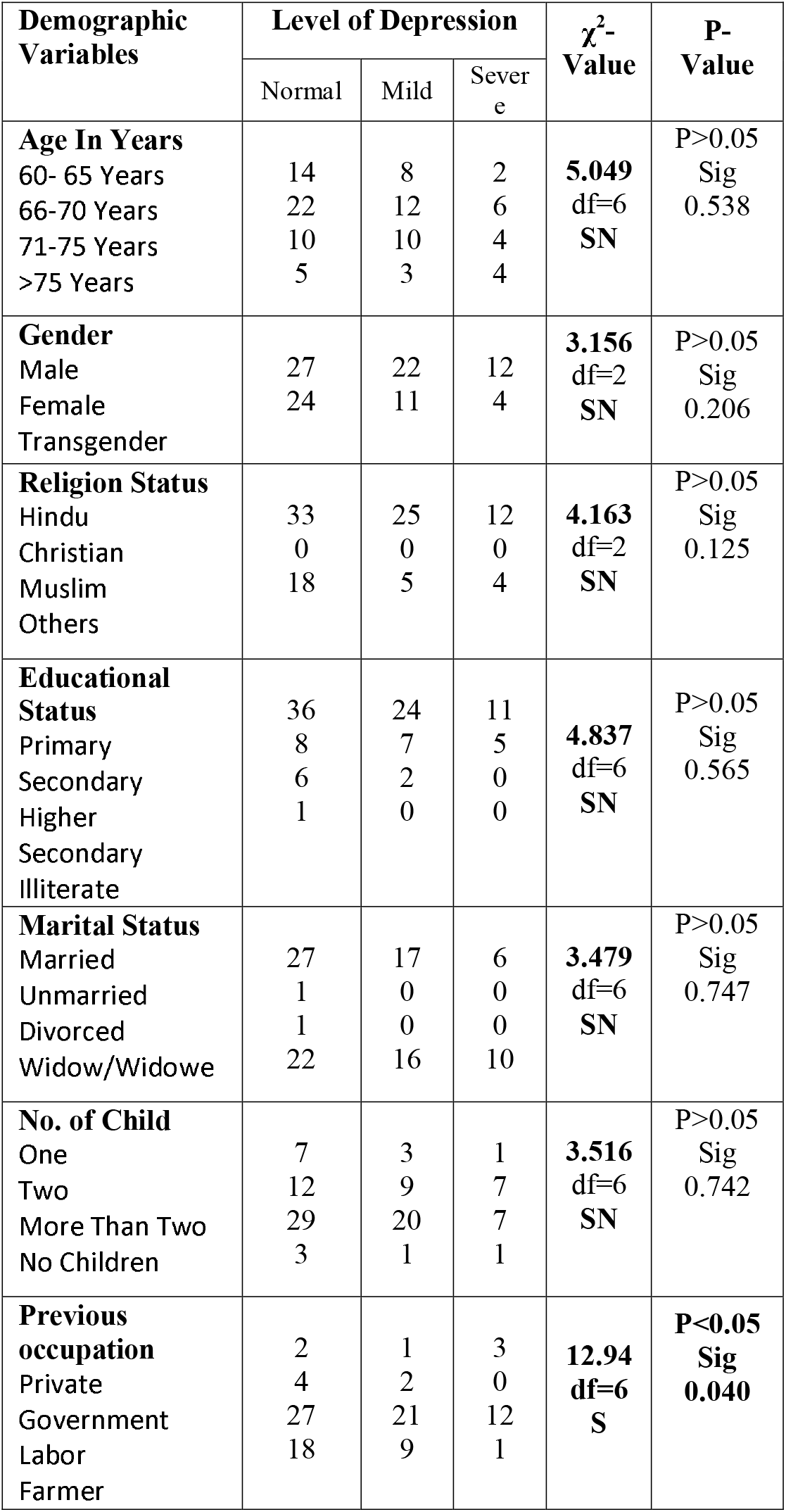
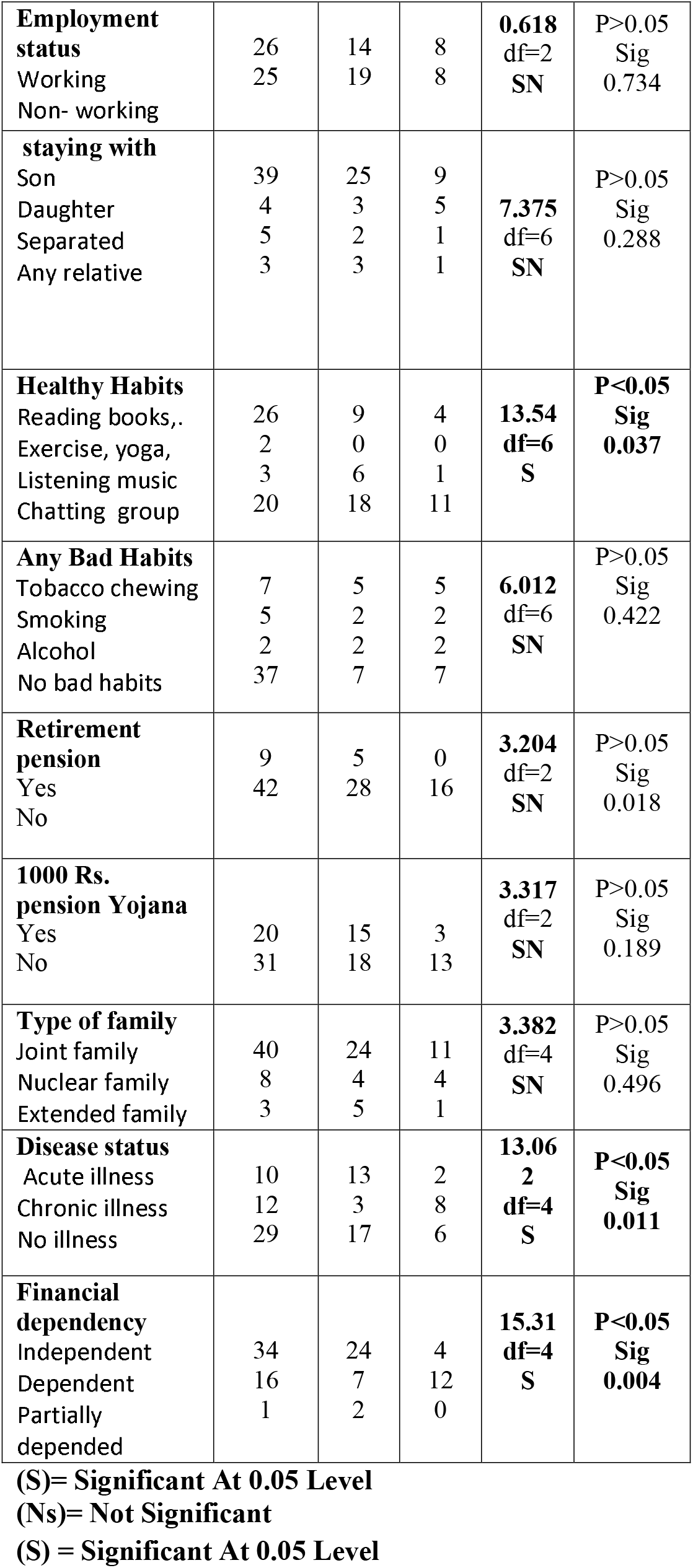
Association between the level of depression with selected demographic variables of geriatric peoples.

### Out of which Previous Occupation, Any Good Habit, Diseases status and Financial dependency of the demographic variable found significant associated at P value <0.05 with Depression level

Hence null hypothesis was rejected and research hypothesis was accepted as above mentioned selected demographic variables **its evidence that there is significant difference between level of depression and geriatric people’s demographic variables**.

## Conclusion

On the basis of analysis of this study the following conclusion were drawn:

The purpose of the present study is to assess the geriatric depression among rural area of Kheda District.

The study consisted of 100 samples that were selected on the basis of Non probability convenient sampling techniques. Based on the objective the data analysis was done. After the assessing the geriatric depression among old age people score was majority had no depression 51(51%), Mild Depression 33 (33%), Severe Depression 16 (16%). This study is in line with previous studies showing the high prevalence of depression in elderly. Results suggest a proper screening for depression among elderly. The high prevalence of depression observed among older adults emphasize on the need of increased community support and availability of health care services for better care of the elderly. There is also an urgent need for greater awareness of depression among family members and community at large. At the same time, it is important to increase community support and create networks for better geriatric care, in accordance with WHO findings.

## Data Availability

All data produced in the present study are available upon reasonable request to the authors

## Ethical Clearance

The study was approved by the institutional ethical committee of Dinsha Patel college of nursing, research committee, there are total 15 members in the committee from various field. **The ethical approval reference number is DPCN-IEC/4062100003** and a formal written permission was gathered from the authority of or Principal of Institute prior to data collection

## Statement of Informed consent

Yes, informed consent form was taken from the participants prior to data collection.

## Acknowledgement

Special thanks to all the participants of the study and Medical officer of the selected PHC for provide us permission for data collection.

